# Trends in Clinical Validation and Usage of Food and Drug Administration (FDA)-Cleared Artificial Intelligence (AI) Algorithms for Medical Imaging

**DOI:** 10.1101/2022.06.19.22276350

**Authors:** Mihir Khunte, Allison Chae, Richard Wang, Rishubh Jain, Yujuan Sun, John R. Sollee, Zhicheng Jiao, Harrison X. Bai

## Abstract

**Objective:** The objective of this study is to examine the current landscape of FDA-approved AI medical imaging devices and identify trends in clinical validation strategy.

**Materials and Methods:** We conducted a retrospective study that analyzed data extracted from the American College of Radiology (ACR) Data Science Institute AI Central database as of November 2021 to identify trends in FDA clearance of AI products related to medical imaging. Product and clinical validation information of each device was gathered from their respective public 510(k) summary or de novo request submission, depending on their type of authorization.

**Results:** Overall, the database included a total of 151 AI algorithms that were cleared by the FDA between 2008 and November 2021. Out of the 151 FDA summaries reviewed, 97 (64.2%) reported the use of clinical data to validate their device. Of these 151 summaries, 81 (53.6%) reported the total number of patient cases used during validation, with the average number of cases being 799 (SD: 1363) and the range of cases spanning from 15 to 9122. A total of 51 (33.8%) AI devices characterized their clinical data as multicenter, 3 (2.0%) as single-center, and the remaining 97 (64.2%) did not specify. The ground truth used for clinical validation was specified in 78 (51.6%) FDA summaries.

**Discussion and Conclusion:** A wide breadth of AI algorithms have been developed for medical imaging. Most of the devices’ FDA summaries mention their use of clinical data and patient cases for device validation, emphasizing their utility in real clinical practice.

## Introduction

Artificial intelligence (AI) is a powerful medical tool that can offer significant value by improving quality of care, decreasing costs, and identifying potential adverse events before they occur.

The large amounts of data being produced in healthcare need to be effectively synthesized and AI presents as an effective tool to help clinicians identify patterns and draw appropriate conclusions from all the information. Medical imaging has been an area of medicine that has seen a lot of innovation with regards to AI. The availability of quantitative data and innovation in deep learning may partially explain the growth of AI applications in this area.

There are few main types of AI products being developed for medical imaging which include computer-aided detection (CADe), computer-aided diagnosis (CADx), computer-aided triage (CADt), computer-aided quantification (CADq), and imaging processing.^1^ The various subgroups not only help describe the main functions being carried out by these AI medical products, but also come with varied levels of risk. For example, the U.S. Food and Drug Administration (FDA) considers CADx devices to be higher risk than CADe.^1^ The level of risk is largely dependent on the amount of human supervision over the algorithm. The FDA has published recommendations for the clinical validation of CADe devices.^2^ The clinical study design and makeup of the clinical validation dataset can impact the safety and effectiveness of the device and introduce potential biases into clinical care. It is critical that these devices undergo thorough clinical validation to ensure it is generalizable to a diverse population and image acquisition landscape. In addition, it is important that the information about the clinical validation process and the study results are publicly available so that clinicians can make informed decisions about which algorithms to introduce in their clinical practice.

To date, more than 150 AI algorithms have been cleared by the FDA for clinical use and the number is only expected to grow with some estimates indicating that the market for AI in medical imaging will grow 10-fold in the next decade.^3^ Questions remain on how these devices were validated and the transparency on the part of manufacturers to share this information. In addition, much is unknown about how the current volume of use in clinical practice. In this study, we examine the current landscape of FDA-approved AI products specifically designed for medical imaging and identify trends in clinical validation strategy as reported in the product FDA summaries. In addition, we identify AI algorithms that have already undergone considerable adoption and dive more deeply into the clinical validation results for this cohort.

## Methods

We conducted a broad, retrospective, and longitudinal study that analyzed data extracted from the American College of Radiology Data Science Institute (ACRDSI) AI Central database. ACRDSI is a professional medical institution that collaborates with radiology professionals, industry leaders, government agencies, patients, and other stakeholders to facilitate the development and implementation of AI applications related to radiology and other imaging domains which maintains a list of U.S. FDA approved AI/ML algorithms. According to the database, there were a total of 151 products that were FDA-cleared as of November 2021. Each product’s information on intended imaging modality, clinical subspecialty, and body region of interest were gathered from the openly accessible database. Data relating to the clinical validation process, such as the use of retrospective patient data, the number of patient cases used in the study, whether the study was performed in a multicenter or single-center institution, and whether ground truth was used, required a more in-depth analysis of their corresponding public 510(k) summary or de novo request submission. In situations where multiple values were provided, values were averaged to provide a holistic measurement of classification or segmentation ability. Statistics on the overall number of product users were collected from the algorithm website, press releases, or third-party articles. Data gathered for each product was verified independently by M.K., A.C., R.W., and R.J. The study does not use private or patient information and therefore did not require oversight by an institutional review board.

## Results

Overall, the database included a total of 151 AI algorithms that were cleared by the FDA between 2008 and November 2021. Of these algorithms, 148 (98.0%) were approved with a 510(k) clearance, with the remaining 3 (2.0%) receiving de novo clearance. Among the 148 devices approved with 510(k) clearance, 52 (35.1%) were based on a primary and/or secondary predicate device from the same company (Figure 1).

**Figure 1:**
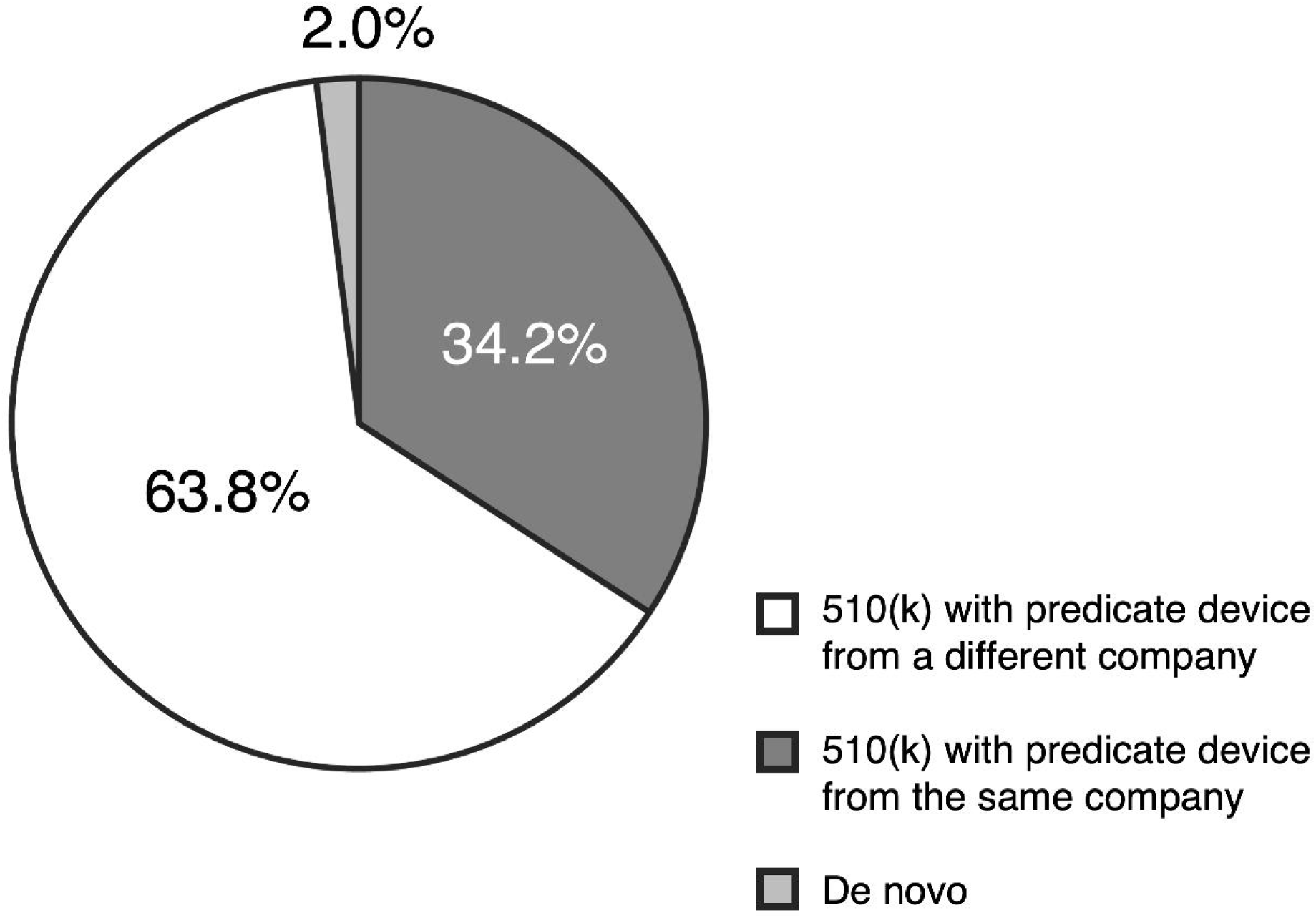
Distribution of FDA-cleared premarket submissions.

CT was the most popular imaging modality (49.0%), followed by MR (25.2%) and X-ray (13.2%). CTA and MAM each constituted less than 3% of the total products. The three most common subspecialty fields that the algorithms were developed for were neuroradiology (33.8%), chest imaging (22.5%), and cardiac imaging (14.6%). Each of the remaining subspecialties constituted less than 7% of the total. Consequently, the most common body regions of interest were brain (32.0%), chest (20.3%), and heart (13.1%). Each of the remaining anatomical regions constituted less than 5% of the total products (Figure 2). These FDA-approved algorithms were developed by companies from 23 different countries, with a majority developed in the USA (49.0%), Israel (10.6%), and France (7.3%).

**Figure 2:**
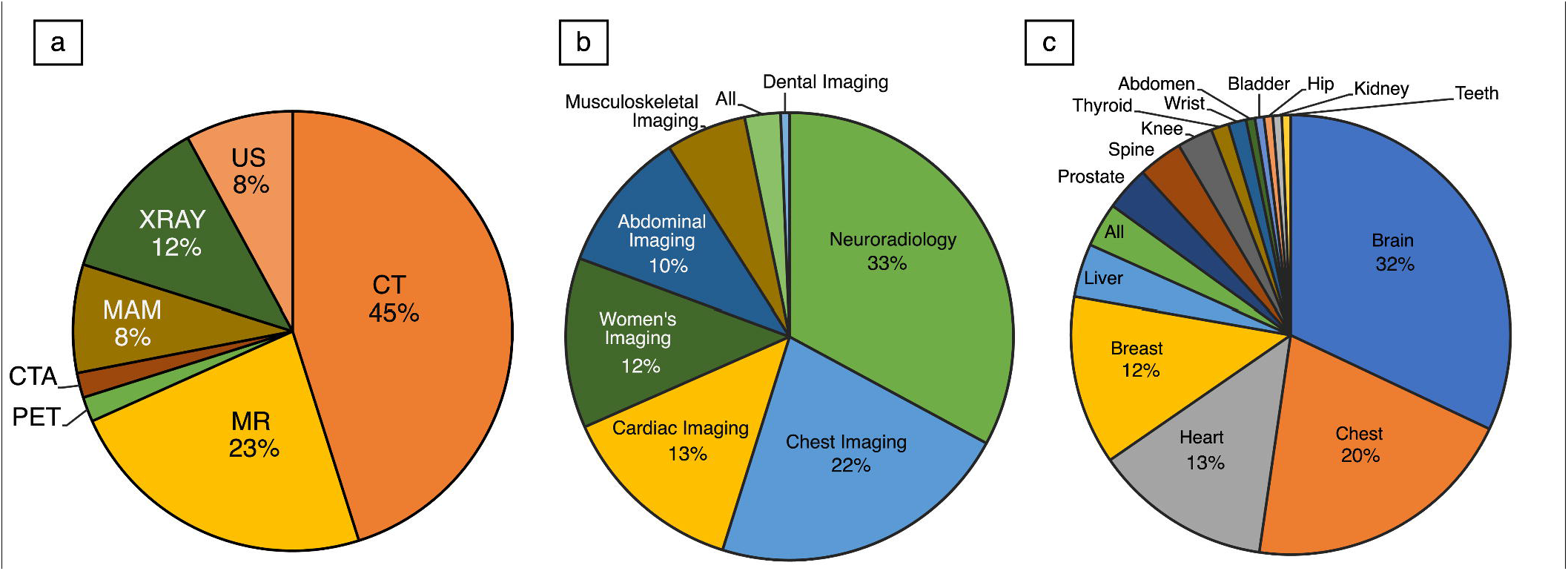
Distribution of imaging modalities, clinical subspecialties, and body regions of interest (a. imaging modalities b. clinical subspecialties c. body regions of interest.).

Out of the 151 FDA summaries reviewed, 97 (64.2%) reported the use of clinical data to validate their device. Of these 151 summaries, 81 (53.6%) reported the total number of patient cases used during validation, with the average number of cases being 799 (SD: 1363) and the range of cases spanning from 15 to 9122. The median value for each of the categories — MAM, XRAY, CT, MR, and US — was 1016, 588, 284, 180, and 378 respectively (Fig. 3). A total of 51 (33.8%) AI devices characterized their clinical data as multicenter, 3 (2.0%) characterized their clinical data as single-center, and the remaining 97 (64.2%) did not specify. The ground truth used for clinical validation was specified in 78 (51.6%) of the FDA summaries, of which 71 were human readers.

**Figure 3:**
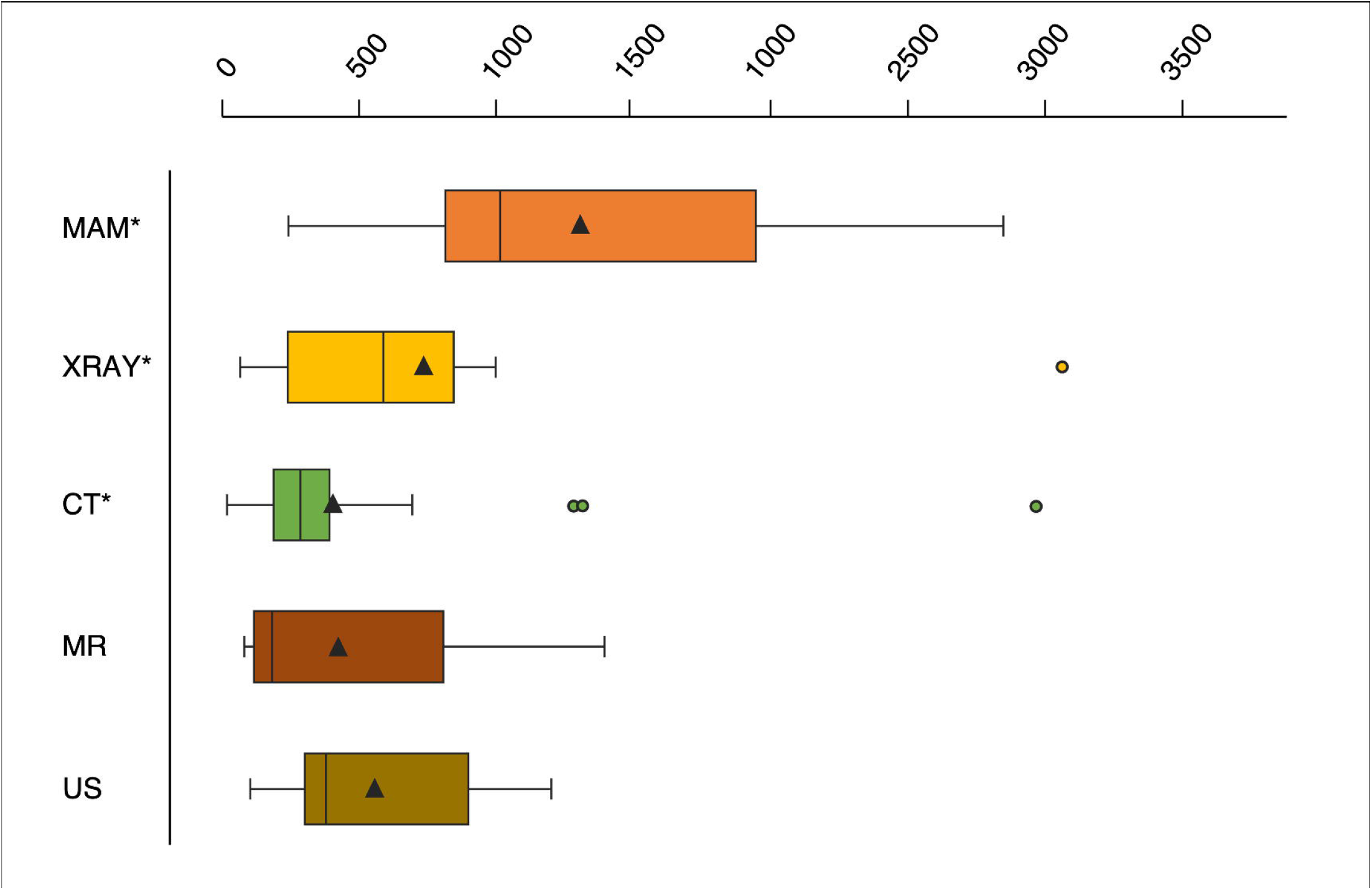
Total number of patient cases used during validation by imaging modality. (MAM: outlier of 9122 is not shown in the graph., XRAY: outlier of 6597 is not shown in the graph., CT: outlier of 4500 is not shown in the graph.) Triangle denotes the mean.

Of the 151 algorithm websites and press releases reviewed, 34 reported some statistics on their number of users (hospitals or other point-of-care centers) at the company or algorithm level. Of 34 algorithms that reported the number of users, 22 had more than 500 users (Table 1). Most of the algorithms (14/22) used CT as their imaging modality with 5 using MR. The algorithms complete a variety of tasks including breast imaging (2), intracranial hemorrhage detection (4), and large vessel occlusion detection (3). Of these FDA summaries analyzed for these algorithms, only 12/22 contained information about the number of patient cases used to validate the algorithm. The median number of cases was 284 with a minimum of 50 cases and a maximum of 2987 cases. Sensitivity and Specificity information was available in the FDA summary for 9/22 algorithms and was greater than 0.85 for both values in all cases. For algorithms with no information about the accuracy listed in the FDA summary, values were identified based on the manufacturer’s website and/or published literature and organized in Table 1.

**Table 1:** Summary information about AI Algorithms with more than 500 public disclosed users.

| Company | Number of Users | Product | Algorithm Function | Modality | Performance Statistics | Number of patients for Clinical Validation |
| --- | --- | --- | --- | --- | --- | --- |
| Volpara Health Technologies | 2000 | Volpara Imaging Software | Provides volumetric assessment of digital x-ray images of the breast. BI-RADS 4th and 5th edition breast density category can be obtained from volumetric breast density. | MAM | No Data Available | No Data Available |
| MeVis Medical Solutions AG | 500+ | Veolity | Provides visualization of chest CT images, measurements of segmented nodules, and CAD software to assist in identification of findings with solid pulmonary nodules features. | CT | Sensitivity = 0.714<br>Specificity = 0.955<sup>10</sup> | 1090<sup>10</sup> |
| ICAD | 2000 | ProFound AI Software V2.1 | Detects malignant soft-tissue densities and calcifications in tomosynthesis images. Marks suspicious soft tissue densities and calcifications. Provides score for regions of interest and cases based on algorithm's confidence they are malignant. | MAM | Sensitivity = 0.915<sup>11</sup><br>Radiologist Specificity with Profound AI = 0.696 vs 0.627 without Profound AI<sup>12</sup> | 1016 |
| Vital Images | 500+ | Vitre CT Brain Perfusion | Calculates cerebral blood flow, cerebral blood volume, local bolus timing, and mean transmit time from CT image data. Aids in visualization of apparent blood perfusion in brain tissues affected by acute stroke and differentiates areas of decreased perfusion. | CT | No Data Available | No Data Available |
| iSchemaView | 1800+ | Rapid ASPECTS | Assesses and characterizes brain tissue abnormalities using CT image data. Segments and analyzes regions of interest to provide ASPECT score which reflects number of regions with early ischemic change per ASPECTS guidelines. | CT | No Data Available | 50 |
|  |  | RAPID ICH | Analyzes non-enhanced head CT images and flags cases with suspicion of ICH. | CT | Sensitivity = 0.899<br>Specificity = 0.943 | 336 |
|  |  | RAPID LVO 1.0 | Analyzes CTA head images and highlights cases with suspected LVO | CTA | Sensitivity = 97%<br>Specificity = 96%<sup>13</sup> | No Data Available |
| Aidoc | 600 | Aidoc Briefcase for iPE Triage | Marks suspected positive cases of Incidental Pulmonary Embolism (iPE) pathologies. | CT | Sensitivity = 0.905<br>Specificity = 0.887 | 268 |
|  |  | Aidoc Briefcase-CSF Triage | Analyzes cervical spine CT images and flags suspected findings of linear latencies in patterns compatible with fractures | CT | Sensitivity = 0.917<br>Specificity = 0.886 | 186 |
|  |  | Briefcase-IFG | Analyzes Abdominal CT images and flags cases with suspected findings of IFG. | CT | Sensitivity = 0.910<br>Specificity = 0.899 | 184 |
|  |  | Aidoc Briefcase-LVO | Analyzes head CTA images and flags suspected cases of LVO | CTA | Sensitivity = 0.888<br>Specificity = 0.872 | 383 |
|  |  | Aidoc Briefcase-ICH and PE Triage | Analyzes non-enhanced head CT images and highlights cases with suspected findings of ICH and PE. | CT | Sensitivity = 0.906<br>Specificity = 0.899 | 2987 |
|  |  | Briefcase-ICH | Analyzes non-enhanced head CT images and highlights cases with suspected findings of ICH. | CT | Sensitivity = 0.936<br>Specificity = 0.923 | 198 |
| TeraRecon | 1300 | Neuro.AI Algorithm | Analyzes brain perfusion from MRI or CT imaging data sets. Displays properties of change in contrast over time and calculates parameters related to brain tissue perfusion and tissue blood volume. | CT, MR | No Data Available | No Data Available |
|  |  | iNtution-Structural Heart Module | Enables visualization and measures of heart structures to assist in pre-operative planning and calcium quantification. | PET, CT, MR | No Data Available | No Data Available |
| Viz. ai | 850 | Viz ICH | Analyzes non-contrast CT images of the head and examines features for indication of intracranial hemorrhage. | CT | Sensitivity = 0.95<br>Specificity = 0.96<br>AUC = 0.97 | 387 |
|  |  | Viz CTP | Analyzes CT perfusion scans of the brain and provides calculations of parameters related to tissue flow and tissue blood volume. | CT | No Data Available | No Data Available |
|  |  | Viz LVO | Analyzes CT angiogram images of the brain and identifies suspected LVO for further review by a physician. | CT | Sensitivity = 0.878<br>Specificity = 0.896<br>AUC = 0.91 | 300 |
| Cortechs.ai | 1000+ | NeuroQuant | Analyzes MR images to automatically label and volumetrically quantify segmented brain structures and lesions. | MR | Sensitivity = 0.688<br>Specificity = 0.904<sup>14</sup> | No Data Available |
|  |  | NeuroQuant MS | Quantitatively analyzes FLAIR lesions and brain structures. | MR | Sensitivity = 0.53<br>Specificity = 0.75<sup>15</sup> | No Data Available |
|  |  | OnQ Prostate | Analyzes MR imaging data to provide automatic prostate quantitation and segmentation. | MR | No Data Available | No Data Available |
|  |  | RSI-MRI+ | Analyzes MR imaging data to provide automatic prostate quantitation and segmentation. | MR | Sensitivity = 0.810<br>Specificity = 0.818<sup>16</sup> | No Data Available |
|  |  | CT CoPilot | Uses CT images of the brain to provide quantification, visuals, and segmentation of structures. | CT | No Data Available | No Data Available |
| DiA Imaging Analysis | 1000 | Lvivo Software Application | Analyzes ultrasounds images and provides segmental evaluation of the left ventricle of the heart. | US | Sensitivity = 0.87<br>Specificity = 0.60<sup>17</sup> | 100 |

## Discussion

The role of AI in healthcare has grown substantially over the past few years. In particular, AI algorithms for use in medical imaging have the potential to greatly improve the efficiency and efficacy of medical professionals worldwide. Despite this, there have been very few studies that provide a broad overview of these FDA-approved AI algorithms. This lack of coverage may contribute to a lack of understanding and/or awareness of these devices in the scientific and medical communities, which may, in turn, lead to their underutilization. Our study aimed to bridge this gap in knowledge by providing an analysis of all current FDA-approved AI algorithms in medical imaging.

As of the end of November 2021, there are a total of 151 unique AI algorithms that have been cleared by the FDA for use in medical imaging. These algorithms were developed by companies based in over twenty different countries, showcasing the global interest in developing and utilizing these technologies. Perhaps intuitively, all twenty-three of these countries were within the top 60 countries ranked worldwide by total GDP.^4^

Importantly, all 151 of the AI devices approved for medical imaging used clinical data to validate their device, with the majority (64.2%) clearly stating so in their FDA summary. Most of these devices characterized their clinical data as multicenter, and, of the algorithms that reported the number, the average number of patient cases used for validation was 799. The average number of patient cases used for validation was highest for AI devices designed for mammography (1016) and ultrasound (588), and lowest for AI devices designed for MR (180). This may be due to the ease and volume at which ultrasound and mammography images can be collected for clinical research compared to MR. This use of large, multicenter clinical data further emphasizes the utility of these AI algorithms in real, clinical practice.

In terms of the imaging modalities that these AI algorithms were designed to augment, CT was the most popular (49%), followed by MR (25.2%). These findings likely reflect the relatively high usage of these two modalities in the healthcare and research communities. For instance, a recent study showed that CT and MR are the two modalities with the most rapidly increasing usage rates in the United States over the past two decades^5^; a separate study showed that CT and MR accounted for more than 50% of new AI research publications in radiology in the past few years.^6^ Similarly, the imaging modality least represented in the list of AI algorithms was PET, likely reflecting how nuclear imaging has been the least involved imaging modality in terms of both healthcare usage rates and research interest.^5, 6^ The most common subspecialty field that the AI algorithms were designed for was the nervous system (33.8%). This is again paralleled by the observation that neuroradiology is the most involved subspecialty in AI research, accounting for roughly one-third of AI-related radiology publications in the past few years. There is ultimately a clear association between the modalities and subspecialties that these AI devices have been designed for and the relative interest/usage of these modalities and subspecialties in the medical community. However, it remains unclear whether the former drove the latter or vice-versa.

A subset of these AI devices provided data on the number of hospitals and other point-of-care centers that used them. Roughly two-thirds of this subset could be considered high-use, with at least 500 users worldwide. Like the observed trends of all 151 devices as previously described, the majority of these high-use AI devices were designed for use with CT (63.6%), followed by MR (22.7%). The respective purposes of these high-use AI devices varied, but the largest proportion was tasked with detecting and qualifying stroke, including intracranial hemorrhage and large vessel occlusion (31.8%). The relatively high global interest in AI devices designed for stroke detection may be due to the rising prevalence of stroke worldwide^4^ and the subsequent demand to better identify it in patients.^7^ As the global patient population continues to age, it is possible that we may see increased interest in AI devices that augment detection of other age-associated diseases, such as ischemic heart disease and cancer.

Notably, one of the high-use AI devices for large vessel occlusion detection, Viz LVO, was recently granted a New Technology Add-on Payment (NTAP) by the Centers for Medicare and Medicaid Services (CMS) in 2021.^8^ This is the first incidence of CMS reimbursement for AI devices used for medical imaging, and it is an important hallmark in clinical medicine. The effect of reimbursement for AI devices is two-fold. Firstly, it incentivizes health care systems in the United States to adopt cost-effective algorithms for clinical use, which will ideally lead to better-optimized systems and patient outcomes. Secondly, it will drive further innovation in AI medical imaging, especially in the area of reimbursed devices. We have already seen a number of AI healthcare companies develop their own stroke detection AI algorithms.^9^

There are several limitations to our study that should be considered. Firstly, we only used data that was made publicly available, such as the ACRDSI AI central database. While the database offers great insight into the current state of AI in healthcare, it is not fully comprehensive, nor does it contain all the information that the companies submitted to the FDA for device approval. For instance, while all 151 companies used clinical data to validate their device, only a subset of companies clearly stated so in their FDA summary. Secondly, there we relatively little data available on the clinical usage of these devices. We ultimately relied on self-reported data from the companies or their users themselves, such as through websites or news articles. This, in turn, may have led to biased results (i.e., companies with more users are more likely to make this information available to the public). These limitations may have hindered us from seeing the full scope of the current AI healthcare landscape.

Ultimately, our results illustrate the wide breadth of AI algorithms that have been developed for medical imaging purposes, with most utilizing clinical data to support their validity for use in real clinical care. Trends in the modalities and subspecialties that these algorithms have been designed for parallel the involvement of these modalities and subspecialties in clinical practice and AI research. Specifically, CT and MR make up the greatest proportion of imaging modalities represented, with neuroimaging like stroke being the most involved subspecialty field. As the intersection between AI and medical imaging continues to grow in importance, it is imperative that physicians recognize and familiarize themselves with these new technologies being developed.

## Data Availability

All data produced in the present study are available upon reasonable request to the authors.

## Data Availability

Collected dataset will be made available from the corresponding author upon reasonable request from other researchers.

## Author Contributions

M.K., A.C., R.W. and H.B. conceived and designed the analysis. M.K., A.C., R.W., and R.J. collected the data. M.K., A.C., R.W., and R.J. performed the analysis. M.K., A.C., R.W., Y.S., and J.S. wrote the paper. Z.J. and H.B. supervised the project. All authors provided critical feedback and helped shape the interpretation of the research.

